# A spatial EHR- and wastewater-informed modeling framework for respiratory virus forecasting under sparse and missing data conditions

**DOI:** 10.64898/2026.05.18.26353485

**Authors:** Lu Zhong, Amanda Bleichrodt, Aakash Pandey, Deborah Kunkel, Lior Rennert

## Abstract

Heterogeneous surveillance data, including electronic health record (EHR) and wastewater, provide complementary information for monitoring infectious disease transmission and healthcare burden. However, surveillance systems often exhibit overlapping spatial gaps, particularly in underserved communities where clinical and environmental data are sparse or unavailable, limiting the performance of existing forecasting methods. Here, we develop a spatial Bayesian renewal framework that integrates heterogeneous surveillance data through a shared latent infection process and mobility-informed spatial interactions, enabling robust disease burden forecasting under incomplete surveillance. We apply the framework to forecast county-level hospital admissions associated with three major respiratory viruses: influenza, SARS-CoV-2 (COVID-19), and respiratory syncytial virus (RSV), across South Carolina. In rolling four-week forecasting horizons, the proposed framework consistently outperforms non-spatial approaches and maintains strong predictive performance even in counties lacking direct wastewater or insufficient EHR observations. By borrowing information across spatially connected regions, the model leverages surveillance-rich communities to improve forecasts in surveillance-poor areas. These results demonstrate that integrating complementary surveillance streams within a spatial Bayesian framework provides a scalable and generalizable approach for infectious disease forecasting and supports public health decision-making where surveillance infrastructure is incomplete.

## Introduction

Reliable forecasting of disease burden is essential for preparing health systems for respiratory disease outbreaks^1^. Electronic health record (EHR)-based surveillance provides timely measures of disease burden and has become an important resource for monitoring respiratory disease dynamics^2^. Recent studies have highlighted the importance of granular forecasting for capturing spatial heterogeneity in disease dynamics and informing local public health decision-making^3–5^. However, EHR-based forecasting can be impacted by unequal healthcare access^6^, incomplete population coverage, reporting delays^7,8^, and regional sparsity in clinical observations^9^. To complement EHR surveillance, alternative surveillance data streams such as wastewater-based epidemiology (WBE) have emerged as promising indicators of community-level transmission^10–14^. Because viral shedding into wastewater occurs independently of healthcare-seeking behavior or diagnostic testing access, wastewater surveillance can capture both symptomatic and asymptomatic infections and often provides earlier signals of disease activity than clinical reporting system^15,16^. Multiple studies have shown that concentrations of viral RNA in wastewater can precede increases in reported cases and hospitalizations for pathogens such as SARS-CoV-2 (COVID) ^17–22^, influenza^23,24^, and respiratory syncytial virus (RSV)^24^.

Despite their complementary strengths^25^, neither EHR- nor wastewater-based surveillance provides completely spatial coverage^2,26^. Wastewater surveillance remains fragmented across the United States, with monitoring available in only 27% of counties and concentrated primarily in urban areas^27^. Likewise, EHR-based surveillance depends on healthcare system participation and patient access to care^28,29^, potentially underrepresenting disease activity in rural and underserved communities^9^. As a result, surveillance gaps in wastewater and clinical reporting systems can co-occur^9,26,27^. Importantly, these gaps are not randomly distributed. In rural and resource-limited communities, wastewater surveillance may be infeasible because centralized sewer infrastructure is limited and septic-system use is common, while comprehensive EHR coverage is often concentrated within large healthcare systems, leaving fragmented clinical signals elsewhere^4,5,30^. These conditions produce structural, rather than merely intermittent, sparsity: some regions lack entire surveillance modalities, such that models that appear reliable at aggregated or data-rich scales may degrade when applied locally^4,31,32^. These surveillance blind spots are especially concerning in communities where delayed access to healthcare and elevated risk of adverse outcomes make timely local forecasting most valuable^3,33^.

Most existing forecasting approaches, including statistical^34–36^, Bayesian^37–39^, machine-learning^40^, and mechanistic models^19^ have been developed for settings with dense and reliable surveillance data or for individual surveillance systems. Their performance often deteriorates when surveillance observations are sparse, heterogeneous, or simultaneously missing across multiple data streams. A key but underexploited opportunity to address this challenge lies in “borrowing strength” across connected regions^41,42^. Human mobility creates epidemiological connections between communities^43–45^, allowing observations from surveillance-rich communities to inform forecasts in surveillance-poor communities. Although previous studies have incorporated spatial information through spatial smoothing^46,47^ or hierarchical models^48,49^, few forecasting frameworks have leveraged mobility-informed spatial connectivity to jointly integrate heterogeneous surveillance streams while accounting for data reliability.

In this study, we develop a spatial Bayesian renewal framework that integrates heterogeneous surveillance observations to forecast hospital admission. The framework extends the Bayesian renewal framework adopted by the CDC^38,39^ for disease forecasting by combining reliability-weighted wastewater measurements, EHR-derived indicators of healthcare burden, and mobility-informed spatial interactions within a unified probabilistic model. By explicitly modelling spatial connectivity, the framework enables information sharing across connected counties and generates robust forecasts even in regions lacking direct wastewater or limited EHR observations. We apply the framework across counties covered by South Carolina’s two largest health systems to forecast hospital admissions for COVID-19, influenza, and RSV across rolling prediction horizons. This work demonstrates how spatially informed integration of heterogeneous surveillance systems can support infectious disease forecasting in underserved and surveillance-poor settings.

## Results

### Surveillance sparsity in South Carolina

To characterize surveillance availability, we first examined the spatial distribution of wastewater (WW) monitoring sites and electronic health record (EHR)-based surveillance across South Carolina (Fig. 1a). Among the state’s 46 counties, only 16 had WW sampling. To assess the availability of clinical data, counties were classified according to hospital admission, with an average of at least one encounter per day considered sufficient for epidemiological analysis. During the study period (July 2024–January 2026), 21 counties met this criterion for COVID-19, 22 for influenza, and only 11 for RSV (Fig. 1b). Integrating WW and EHR surveillance revealed substantial spatial gaps in surveillance coverage. For COVID-19, only 10 counties had both WW monitoring and sufficient hospital admission data, whereas 19 counties had neither surveillance source. Similar patterns were observed for influenza (10 counties with both surveillance systems and 18 with neither) and RSV (6 counties with both surveillance systems and 25 with neither). These findings reveal extensive spatial surveillance blind spots, particularly for RSV, where more than half of counties lacked both environmental and clinical surveillance.

**Figure 1.**
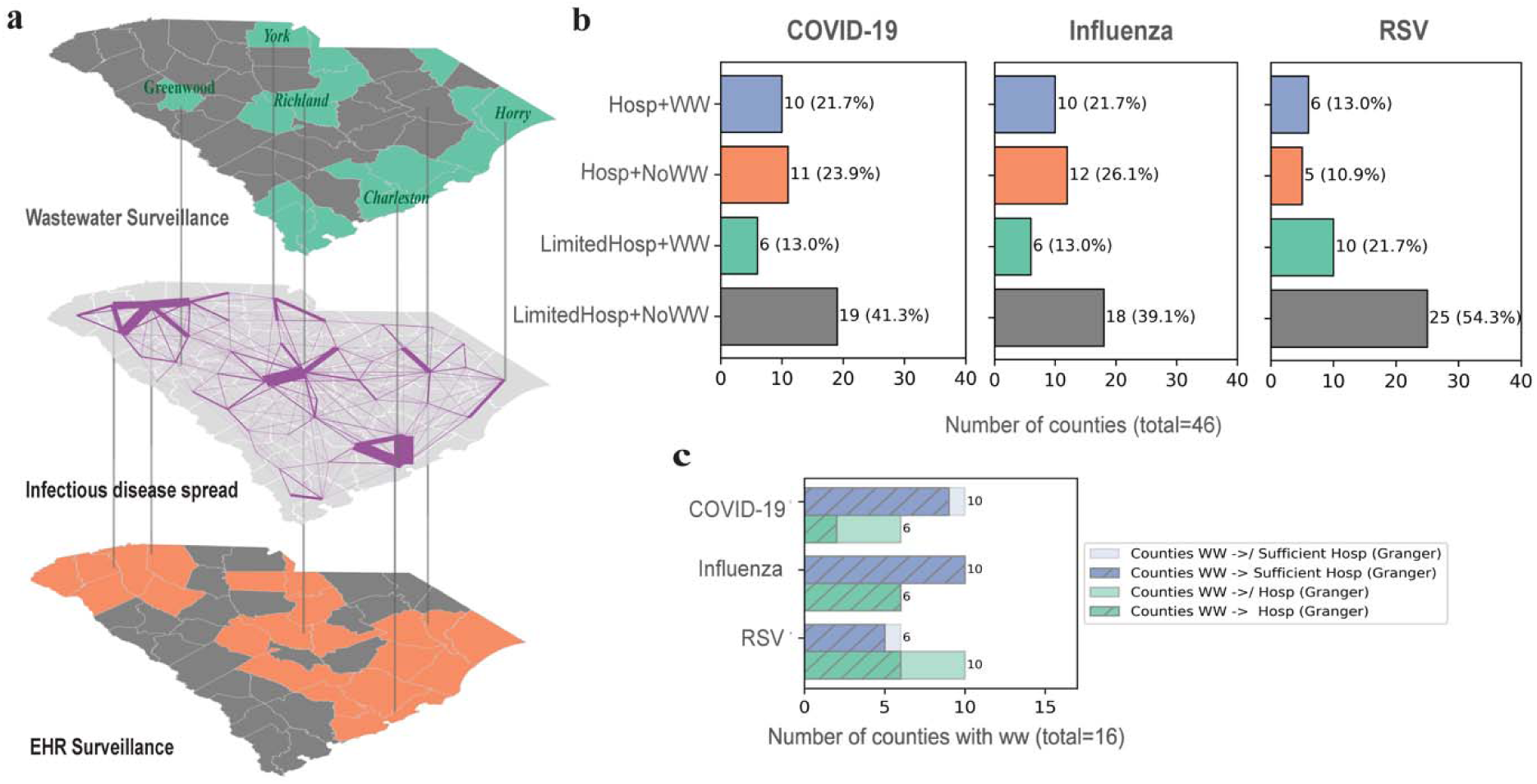
Spatially heterogeneity of wastewater and EHR surveillance across South Carolina. (a) Geographic distribution of wastewater (WW) surveillance sites and electronic health record (EHR) surveillance coverage across South Carolina counties. The top panel identifies counties with active WW monitoring. The middle panel depicts the inter-county mobility network, representing potential pathways for infectious disease transmission. The bottom panel illustrates the average daily hospital admission from Prisma Health and MUSC by county; counties averaging more than one encounter per day are considered to have sufficient clinical surveillance data and are highlighted in orange. **(b)** Distribution of counties across four surveillance contexts for COVID-19, influenza, and RSV: counties with both surveillance sources, WW only, EHR only, or neither. **(c)** Number of counties in which WW viral concentrations significantly Granger-caused hospital admissions (*P* < 0.05), stratified by whether counties had sufficient hospital admissions for epidemiological analysis. These results reveal pronounced spatial disparities in surveillance infrastructure and demonstrate that wastewater–clinical associations differ across monitored regions.

To assess whether WW surveillance provides information beyond EHR surveillance, we evaluated the temporal association between wastewater viral concentrations and hospital admissions using Granger causality analysis (Fig. 1c). Significant Granger-causal relationships (*P* < 0.05) were identified in 11 counties for COVID-19, 16 for influenza, and 11 for RSV. However, the lead time between wastewater signals and hospital admissions varied considerably across counties and pathogens (Table S1), indicating substantial spatial heterogeneity in the temporal relationship between environmental and clinical surveillance. These findings suggest that wastewater surveillance provides additional information for disease monitoring, while its predictive value varies across regions.

### Spatial Bayesian renewal framework enables spatial information borrowing

To address incomplete surveillance, we developed a spatial Bayesian renewal framework that integrates wastewater (WW) surveillance, electronic health record (EHR)-based hospital admissions, and mobility-informed spatial interactions (Fig. 2 and see Methods). In this framework, wastewater measurements provide reliability-weighted information about local transmission intensity, while hospital admissions provide delayed observations of the underlying infection burden. The mobility matrix links these county-specific latent processes by allowing transmission in one county to depend partly on infections in connected counties. Therefore, counties with richer WW or EHR observations help constrain the shared spatial infection process, and this information propagates to sparsely monitored or unmonitored counties through the mobility-weighted transmission structure. This is the mechanism that enables spatial borrowing strength.

**Figure 2.**
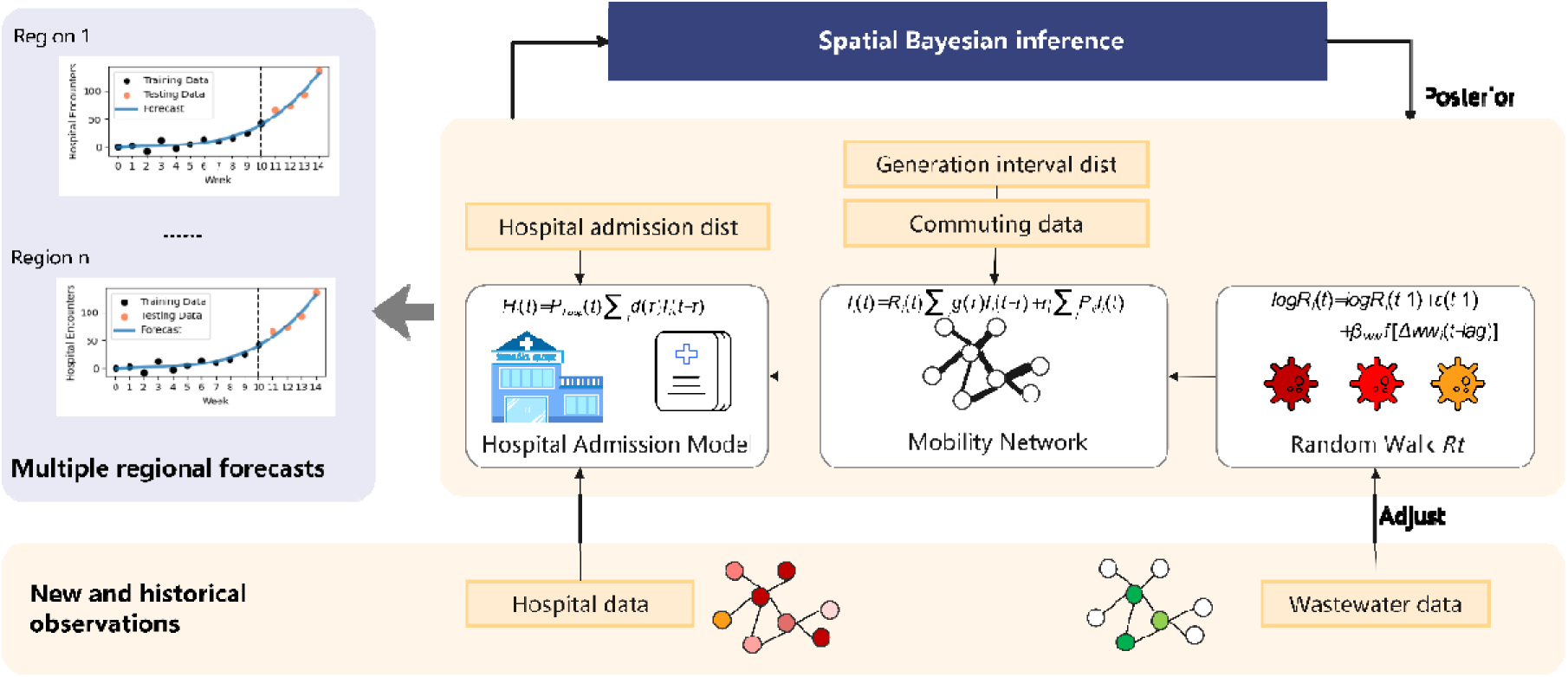
Schematic overview of the Spatial Bayesian renewal framework. The framework integrate heterogeneous surveillance data through a shared latent infection process. Wastewater measurements inform the temporal evolution of the effective reproduction number (), while a mobility-derived spatial transmission network () enables information sharing across counties. The resulting latent infections are linked to observed hospital admissions () through an observation model, allowing regional forecasting under incomplete surveillance.

We evaluated the framework using rolling-origin forecasts of weekly county-level hospital encounters over forecasting horizons of 1–4 weeks. At each prediction date, models were trained on the preceding 12 weeks of surveillance data and evaluated on subsequent observations from September 3, 2024, to December 30, 2025. To quantify the contributions of spatial information borrowing and wastewater surveillance, we compared the proposed framework with the original Bayesian renewal model^38,39^, each implemented with and without wastewater observations. Model performance was assessed separately in four surveillance settings defined by the sufficiency of hospital admissions (HOSP) and WW data: (Group 1) counties with sufficient HOSP and WW data, (Group 2) counties with sufficient HOSP but no WW data, (Group 3) counties with limited HOSP but available WW data, and (Group 4) counties with limited HOSP and no WW data. Forecast accuracy was evaluated by percentage agreement (PA), root mean square error (RMSE), and weighted interval score (WIS).

### Spatial borrowing improves forecasting

Table 1 and Figure 3 summarize forecast performance for counties stratified by the availability of sufficient hospital (HOSP) admission data and wastewater (WW) data. Taking the COVID-19 as an example, among counties with HOSP data (Groups 1 and 2), the ARIMA model achieved the highest performance at short forecast horizons, with 1-week-ahead accuracy reaching 0.867 in Group 1 and 0.875 in Group 2. However, its performance declined more rapidly with increasing forecast horizons. The Spatial Bayesian Renewal model consistently outperformed the original (non-spatial) Bayesian Renewal model across all forecast horizons. For example, at the 1-week horizon, performance improved from 0.766 to 0.813 in Group 1 and from 0.764 to 0.825 in Group 2, with similar improvements observed at 2-, 3-, and 4-week horizons. Performance also varied according to data availability. In counties without sufficient HOSP data (Groups 3 and 4), ARIMA and the non-spatial Bayesian Renewal model could not be applied, because ARIMA generated implausible negative predictions, and the non-spatial Bayesian model could not be adequately estimated from the limited data. Nevertheless, the spatial model maintained moderate and stable performance across forecast horizons (0.654–0.671 in Group 3 and 0.681–0.697 in Group 4). These findings demonstrate that spatial coupling enables robust forecasting even when local surveillance data are sparse or unavailable.

**Figure 3.**
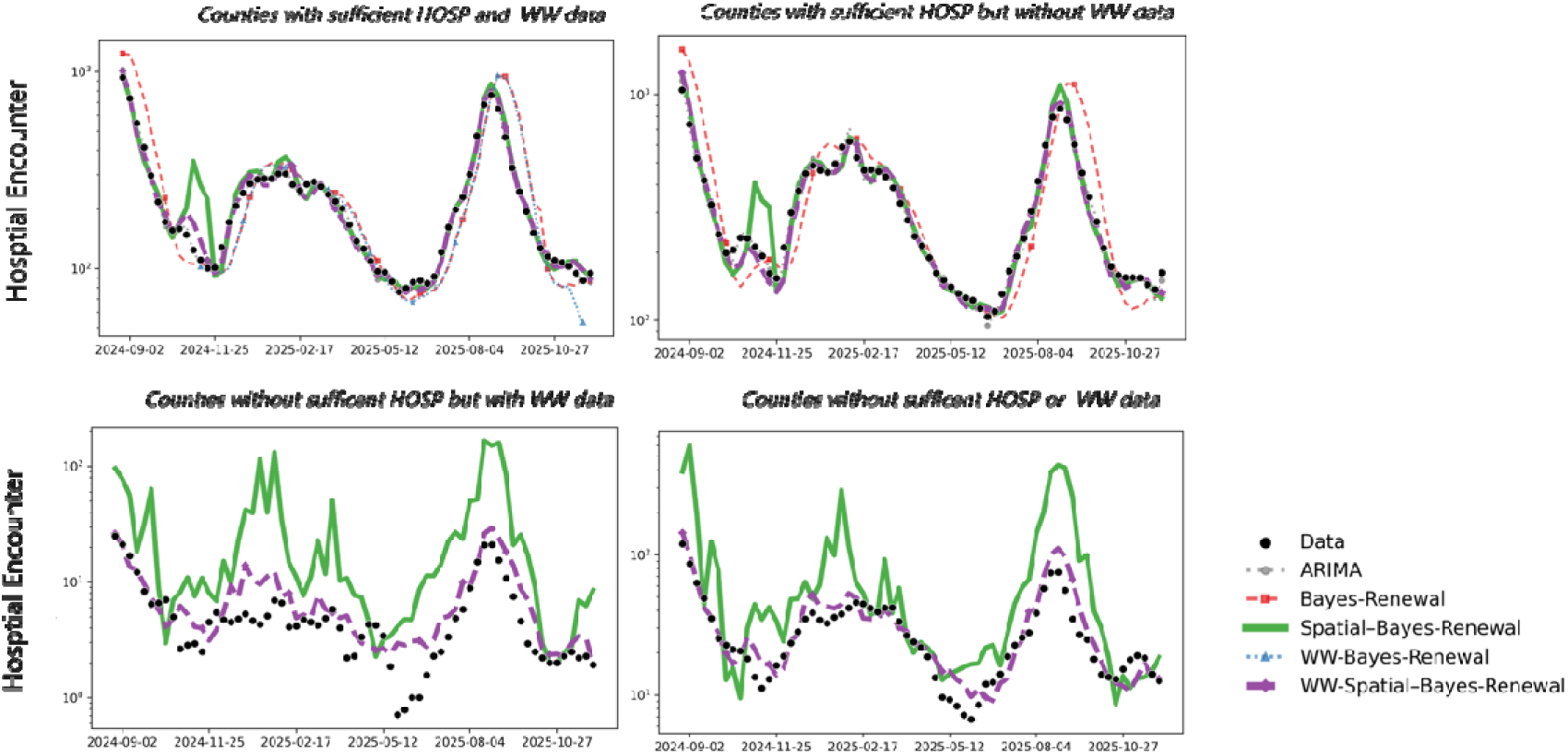
County-level forecasts of COVID-19 hospital admissions under different surveillance settings. Counties are grouped according to the availability of hospital admission (HOSP) and wastewater (WW) surveillance: (1) sufficient HOSP data with WW data, (2) sufficient HOSP data without WW data, (3) limited HOSP data with WW data, and (4) limited HOSP data without WW data. Observed hospital admissions are compared with forecasts generated by ARIMA, the Bayesian renewal model, the wastewater-informed Bayesian renewal model, the proposed spatial Bayesian renewal model, and the proposed spatial wastewater-informed Bayesian renewal model.

**Table 1.**
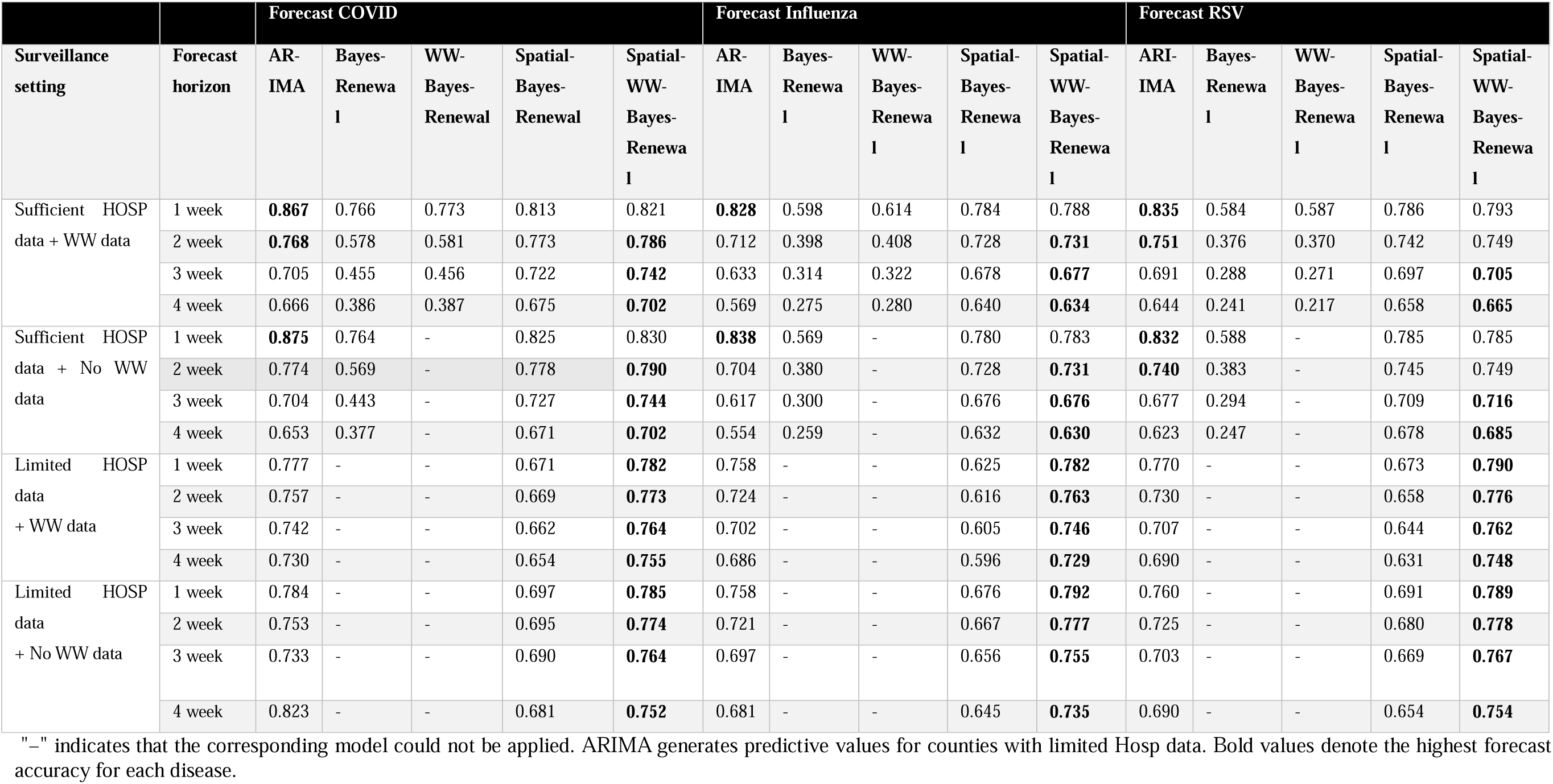
Comparison of forecasting accuracy (percentage agreement) for county-level hospital admissions using spatial and non-spatial forecasting models. Forecast performance is evaluated for COVID-19, influenza, and RSV under four surveillance settings and forecasting horizons ranging from 1 to 4 weeks ahead.

Similar improvements were observed for influenza and respiratory syncytial virus (RSV). Across both pathogens, the spatial Bayesian renewal model consistently achieved higher and more stable forecasting accuracy than its non-spatial counterpart, particularly in counties with limited hospital surveillance. Consistent results were obtained using RMSE and Weighted Interval Score (WIS) (Tables S3–S4).

### Wastewater surveillance further improves forecasting

To evaluate the contribution of wastewater (WW) surveillance, we compared forecasting performance with and without WW observations. Across all surveillance settings, incorporating WW data generally improved forecasting accuracy, with the largest gains observed in the proposed spatial Bayesian renewal model (Table 1). For COVID-19, one-week-ahead forecasting accuracy increased from 0.813 to 0.821 in Group 1 and from 0.825 to 0.830 in Group 2 after incorporating WW data. The greatest improvements were observed in counties with limited hospital admission (HOSP) data (Groups 3 and 4), where forecasting accuracy increased from 0.671 to 0.782 and from 0.694 to 0.785, respectively. Similar improvements were observed for influenza and respiratory syncytial virus (RSV).

Figure 3 illustrates representative one-week-ahead forecasts for COVID-19 across the four surveillance settings. In counties with sufficient HOSP data (Groups 1 and 2), all models captured the overall epidemic trajectory, although the non-spatial Bayesian renewal model tended to miss epidemic peaks. The spatial models more accurately reproduced both the timing and magnitude of observed hospital admissions. In surveillance-limited counties (Groups 3 and 4), the spatial model maintained stable forecasts despite limited clinical observations, and incorporating WW data further improved agreement with the observed trajectories. Similar patterns were observed for influenza and RSV (Figs. S1–S2). All the results indicate that spatial information borrowing provides the primary improvement in forecasting performance, whereas wastewater surveillance contributes complementary gains, particularly in regions with limited hospital surveillance.

### Forecasting gains vary across regional surveillance contexts

Figure 4 illustrates the spatial distribution of forecast accuracy across counties for COVID-19 (See Figure S3-S4 for the results for Influenza and RSV). Among counties with sufficient HOSP data, the spatial model shows noticeable improvements compared with the non-spatial model. The largest gains are observed in counties such as Greenville (accuracy increasing from 0.79 to 0.90) and Richland (from 0.77 to 0.91), which function as major population and commuting hubs with richer data availability. These counties likely benefit more from spatial information sharing because they are strongly connected to surrounding areas through mobility patterns and healthcare utilization networks. In contrast, counties such as Georgetown (accuracy decreasing slightly from 0.80 to 0.78) and Horry (accuracy increasing modestly from 0.80 to 0.82) show minimal improvement. This suggests that in coastal areas, where local transmission dynamics may be less influenced by neighboring counties, spatial borrowing contributes less additional predictive value. For counties with insufficient HOSP data, where the non-spatial model could not generate reliable forecasts, the Spatial Bayesian Renewal model nonetheless achieved good predictive performance. The highest forecast accuracies were observed in counties such as Darlington (≈0.797), Cherokee (≈0.797), and Lee (≈0.795), all of which are inland counties located near highly connected hub regions. These spatial patterns suggest that the effectiveness of information borrowing depends on regional connectivity, with highly connected counties benefiting more from neighboring surveillance information than relatively isolated regions.

**Figure 4.**
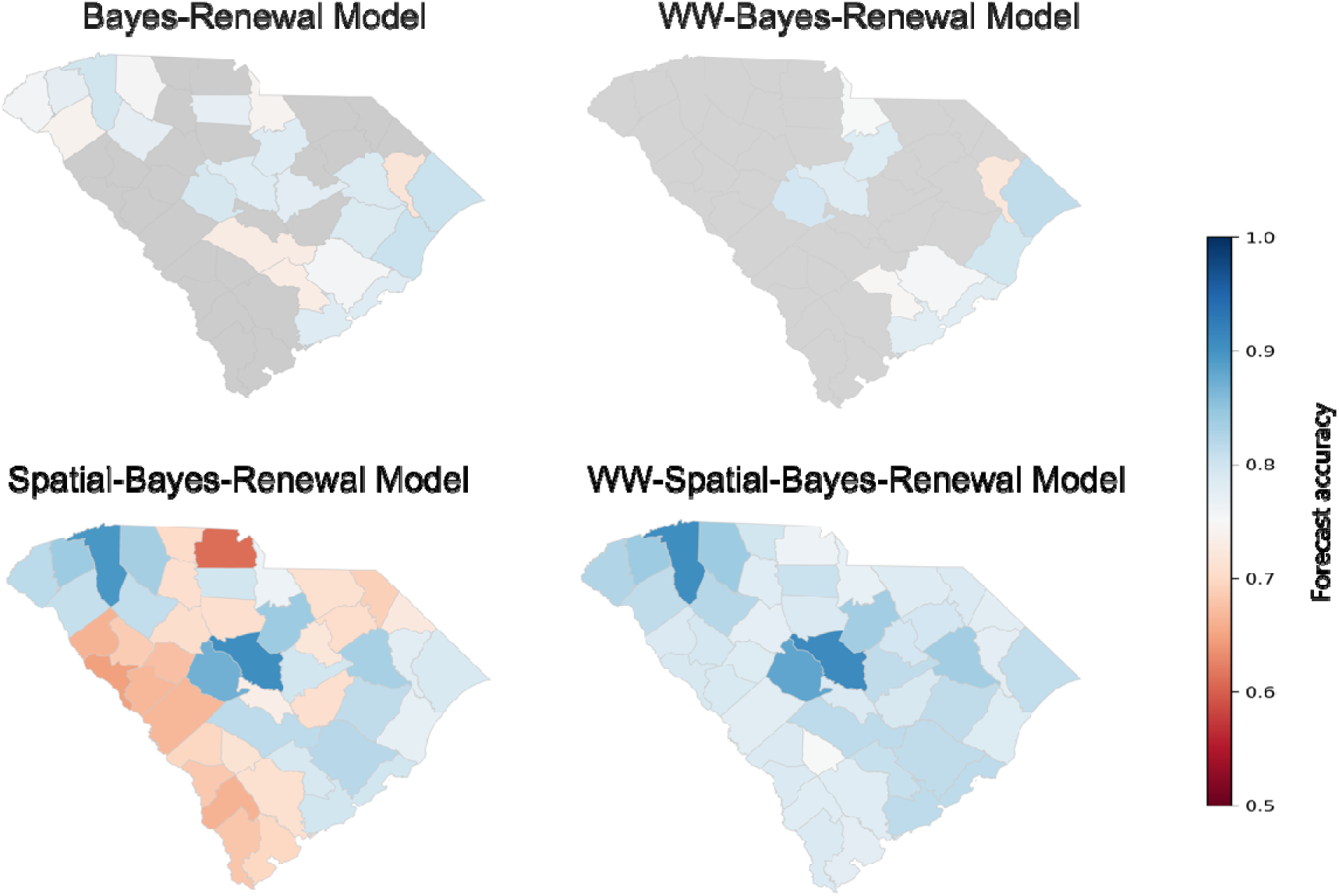
Spatial variation of forecast accuracy for COVID-19 across SC counties. Counties are colored according to forecasting accuracy in one week ahead for each model, with darker blue indicating higher accuracy and red indicating lower accuracy.

## Discussion

In this study, we developed a spatial Bayesian renewal framework that extends the CDC-adopted Bayesian renewal modeling approach by integrating wastewater surveillance, EHR-based hospital admission, and mobility-informed spatial interactions, enabling information borrowing across counties to improve prediction of hospital encounters for multiple respiratory pathogens. Across three major respiratory pathogens, i.e., SARS-CoV-2, influenza, and RSV, the spatial framework consistently improved forecast accuracy compared with non-spatial models. These improvements were driven by two complementary features. First, mobility-informed spatial coupling enabled information borrowing across connected counties, strengthening inference in regions with sparse surveillance data. Second, wastewater measurements provided early signals of local transmission, further improving forecast accuracy before changes were reflected in hospital admissions. Importantly, the framework extends the utility of wastewater surveillance beyond monitored locations by propagating epidemiological information through mobility-connected regions, thereby improving forecasting in counties with limited or no wastewater observations. These findings have important implications for public health surveillance in settings with limited data availability, especially for rural and underserved regions in the US ^50^ and many low- and middle-income countries, where either wastewater infrastructure is limited or absent and clinical surveillance systems are often fragmented^11,51^.

Prior approaches to incomplete surveillance have included imputing missing values before prediction, explicitly encoding missingness or time since observation, adjusting recent observations for reporting delays, and stabilizing local estimates through spatial smoothing or hierarchical borrowing. However, each address only part of the problem created by simultaneous sparsity across time, location, and surveillance modality. Imputation is particularly problematic when a data stream is structurally absent rather than intermittently missing. For example, imputing EHR observations for a county requires assumptions about that county’s health-system coverage, healthcare access, and care-seeking patterns; likewise, imputing wastewater RNA concentrations requires assumptions about sewer connectivity and the population represented by a monitored sewer shed, and may be indefensible in communities primarily served by septic systems^5,52,53^.Thus, imputed values may reflect assumed surveillance infrastructure rather than underlying disease activity, potentially conflating true epidemiologic differences with differences in observation. Missingness-aware and delay-adjustment methods improve the treatment of irregular or delayed measurements but do not themselves recover information for regions lacking multiple local signals, while conventional spatial pooling may rely on generic adjacency or exchangeability rather than epidemiologic connectivity. In contrast, our framework does not reconstruct unobserved surveillance streams as though they had been measured. Instead, it links reliability-weighted wastewater and hospital observations to a shared latent infection process, propagates information over time through the renewal model, and borrows information across counties through mobility-informed (spatial) transmission. Consequently, observations from surveillance-rich counties constrain latent infections and hospital burden in connected surveillance-poor counties, with uncertainty carried through the Bayesian posterior predictive distribution, enabling stable forecasts when local observations are temporally sparse or entirely unavailable.

Furthermore, our results demonstrate that the benefits of spatial borrowing depend on regional location. Counties near major population and mobility hubs can substantially improve forecast accuracy by leveraging information from neighboring regions, whereas spatially isolated counties, particularly along the coast, benefit much less. These findings highlight that spatial connectivity influences the effectiveness of regional forecasting and suggest that surveillance resources may be deployed more efficiently by prioritizing wastewater monitoring in spatially isolated regions while leveraging information sharing among highly connected counties. Such strategies could improve regional situational awareness, healthcare demand forecasting, and preparedness for future respiratory disease outbreaks ^54–56^.

Despite these advantages, several limitations should be considered. First, the mobility matrix used in this study is derived from commuting data and may not fully capture temporal changes in movement patterns. Second, wastewater measurements remain subject to laboratory variability, environmental influences, and sampling inconsistencies; improvements in measurement standardization and data processing could further enhance forecasting accuracy. Third, the assumptions regarding hospitalization delay distributions may vary across pathogens, demographic groups, and time, which could influence inference. Although the Bayesian framework partially accounts for these uncertainties, model performance still depends on the validity of these structural assumptions.

The increasing availability of wastewater surveillance presents new opportunities for regional infectious disease forecasting. Our results indicate that explicitly modeling spatial coupling and accounting for variability in wastewater signal reliability leads to more stable regional inference than treating surveillance sites independently. Broadly, this approach advances wastewater surveillance from isolated site-level monitoring toward a coordinated regional early-warning system^57,58^ providing a scalable strategy for routine infectious disease surveillance and future pandemic preparedness^59^.

## Methods

### Dataset

Three datasets were used in this study: electronic health record (EHR)-based hospital admissions, wastewater (WW) surveillance, and inter-county mobility data.

### Electronic health record (EHR) data

The prediction target was weekly county-level hospital admissions associated with COVID-19, influenza, and respiratory syncytial virus (RSV). Hospital admission data were obtained from Prisma Health and the Medical University of South Carolina (MUSC), the two largest healthcare systems in South Carolina. These systems provide clinical surveillance covering all 46 counties in the state. Weekly hospital admissions were used as the primary outcome for model training and evaluation.

### Wastewater (WW) surveillance data

Wastewater surveillance data were obtained from the Centers for Disease Control and Prevention (CDC) National Wastewater Surveillance System (NWSS) ^60^. County-level viral concentration measurements were collected for SARS-CoV-2, influenza, and RSV and aggregated to weekly resolution to align temporally with the hospital admission data.

### Mobility data

Inter-county mobility data were derived from commuting flows reported in the 2015–2019 American Community Survey (ACS) conducted by the U.S. Census Bureau^61^. The commuting network was used to construct a normalized mobility matrix describing population movement between counties, which was incorporated into the spatial transmission model to represent epidemiological connectivity.

### Preprocess

To ensure consistency across datasets and enable robust temporal analysis, several preprocessing steps were applied. (1) Temporal resolution Adjustment: All datasets were harmonized to daily frequency for training and testing. For hospital admission data originally reported on a weekly basis, daily averages were computed to distribute weekly totals evenly across corresponding days. For wastewater surveillance data, spline interpolation was employed to generate smooth daily estimates from the original reporting intervals, preserving temporal trends while minimizing discontinuities. (2) Normalization of wastewater: For counties with multiple wastewaters monitoring sites, viral concentrations were averaged to obtain a county-level estimate; for counties with a single monitoring site, the reported concentration was used directly. County-level viral concentrations were then log-transformed to stabilize variance and reduce the influence of extreme observations. The model was trained on daily data, and daily forecasts were aggregated to weekly hospital admissions for evaluation.

### Granger causality tests

To assess whether wastewater signals provide leading information for hospital admissions, we performed Granger causality test^62^. Weekly wastewater viral concentrations and hospital admission counts were evaluated over lag periods of up to four weeks. For each lag, the test examined whether incorporating past wastewater measurements significantly improved prediction of current hospital admissions compared with a model using past hospital admissions alone. A *P* value < 0.05 was considered evidence of a statistically significant Granger-causal relationship at the corresponding lag.

### Spatial Bayesian renewal Framework

We extend the classical Bayesian renewal framework by incorporating wastewater surveillance and spatial interactions into a shared latent infection process. The effective reproduction number in county *i*. evolves according to a stochastic random walk on athel go scale,

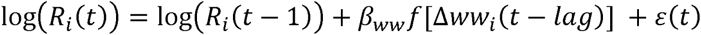

Where *ε*(*i*) is the stochastic term. *β_WW_* quantified the effect of wastewater signals on transmission, and Δ*WW*(*t* − *lag*) represents the change of wastewater viral concentration at a specific lag. The activation function

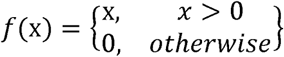

ensures that only increasing wastewater signals contribute positively to transmission intensity, enabling wastewater to act as an early-warning driver of rising infections while preventing spurious decreases in noisy measurements from artificially suppressing transmission estimates. For counties without wastewater surveillance, *f*(x) =0. The wastewater-informed reproduction number drives a renewal process that generates latent infections,

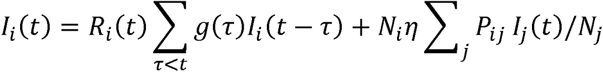

Where g(*τ*) is the generation interval distribution, *P_ij_* is the normalized mobility matrix describing movement from county *i*, to county *j* The coefficient *η* controls the strength of spatial interactions. The notation *N_i_* represent the total population at county *i*. The first term represents local transmission, whereas the second term captures infection importation from spatially connected counties. Expected hospital admissions are linked to latent infections through the hospitalization delay distribution,

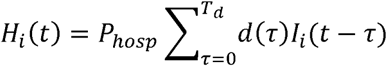

where *d*(*τ*) represents the infection-to-hospitalization delay distribution, *P*_hosp_ (*t*) is the probability of hospitalization given infection. Observed hospital admissions are modeled using a Negative Binomial likelihood,

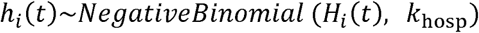

where *k*_hosp_ is the overdispersion parameter.

### Model estimation with partially observed regions

In many real-world surveillance systems, only a subset of locations has monitoring data on wastewater and hospital admissions for training. Let *O* denote the set of observed regions and *u* as the set of un-observed region. The model parameters *Θ* {*β_WW_*, η, *P*_osp_, *k*_hosp_, σ} are inferred using the observed data from regions in *O*. The corresponding posterior distribution of the parameters is given by:

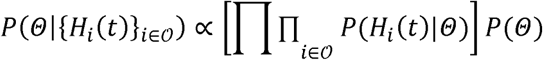

where *H_i_*(*t*) represent the hospital admission observations, respectively, for each observed county *i* ϵ *O*. Counties without direct observations (*i* ϵ *u*) do not contribute a likelihood term, but their latent infection trajectories *I_i_*(*t*) are still inferred through spatial coupling in the renewal process. After posterior inference, forecasts are generated for both observed and unobserved regions using the posterior predictive distribution:

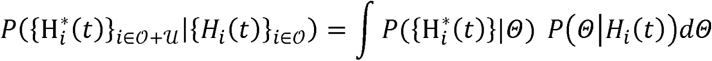

where H*_i_*^*^(*t*) denotes posterior predictive hospital admissions. Posterior inference was performed using Hamiltonian Monte Carlo with the No-U-Turn Sampler (NUTS)^54^ as implemented in NumPyro. Four Markov chains were run, each with 100 warm-up iterations followed by 100 posterior samples.

### Experimental Setup

The prediction target was weekly county-level hospital admissions for COVID-19, influenza, and RSV. Model inputs consisted of wastewater viral concentrations, hospital admission data, county-level mobility data, and fixed epidemiological distributions for the generation interval, *g*(*τ*), and the infection-to-hospitalization delay, *d*(*τ*). Forecasting performance was evaluated using a rolling-origin design. For each prediction date, the preceding 12 weeks of surveillance data were used as the training set, and hospital admissions during the following 1–4 weeks served as the test set. The training and testing procedure were repeated weekly from September 3, 2024, to December 30, 2025, providing a comprehensive evaluation across different epidemic phases.

### Model Comparison

The proposed spatial Bayesian renewal framework was compared with three benchmark models: ARIMA, the original Bayesian renewal model^38,39^, and a wastewater-informed Bayesian renewal model (without spatial coupling). All models were evaluated under the same experimental setup, using identical training and testing datasets, rolling-origin forecasting design, forecast horizons (1–4 weeks), and evaluation metrics (PA, RMSE, and WIS), ensuring a fair comparison of forecasting performance. Percentage Agreement (PA) measures the agreement between predicted and observed hospital admissions and is defined as

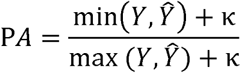

where *Y* and *Ŷ* denotes the observed and predicted hospital admissions, respectively. The constant κ is introduced to stabilize the metric when admission counts are zero or very small, thereby preventing disproportionately large fluctuations in PA. In this study, κ was set to 5. PA ranges from 0 to 1, with larger values indicating better agreement. RMSE measures the average magnitude of prediction errors, with lower values indicating higher forecasting accuracy. WIS evaluates probabilistic forecast performance by jointly assessing calibration and sharpness of the prediction intervals, with lower values indicating better probabilistic forecasts.

## Data Availability

The authors do not have permission to share data.

## Supplementary Information

**Table S1.** Summary of counties with available wastewater (WW) datasets and their Granger causality relationships with hospital admissions. Among the 46 counties in South Carolina, 16 have wastewater data available from July 2024 to 2025. The reported lag represents the number of weeks by which wastewater signals precede hospital admissions with the strongest predictive association. The p-value corresponds to the Granger causality test, with smaller values indicating stronger evidence that wastewater data predict subsequent hospital admissions.

| Counties | pop | WW data (COVID) | WW data (Influenza) | WW data (RSV) |
| --- | --- | --- | --- | --- |
| Beaufort | 187098 | $lag = 2$ [$P = 0.0408$] | $lag = 3$ [$P = 0.0001$] | $lag = 2$ [$P = 0.7997$] |
| Berkeley | 229781 | $lag = 3$ [$P = 0.0050$] | $lag = 3$ [$P = 0.0028$] | $lag = 4$ [$P = 0.0049$] |
| Charleston | 408530 | $lag = 1$ [$P = 0.0001$] | $lag = 3$ [$P = 0.0001$] | $lag = 4$ [$P = 0.0343$] |
| Dorchester | 161331 | $lag = 3$ [$P = 0.0001$] | $lag = 3$ [$P = 0.0001$] | $lag = 1$ [$P = 0.1501$] |
| Georgetown | 63406 | $lag = 1$ [$P = 0.0072$] | $lag = 3$ [$P = 0.0001$] | $lag = 2$ [$P = 0.1228$] |
| Greenwood | 69351 | $lag = 3$ [$P = 0.1030$] | $lag = 1$ [$P = 0.0002$] | $lag = 2$ [$P = 0.0400$] |
| Hampton | 18565 | $lag = 1$ [$P = 0.1665$] | $lag = 1$ [$P = 0.0005$] | $lag = 2$ [$P = 0.2186$] |
| Horry | 351038 | $lag = 1$ [$P = 0.1573$] | $lag = 1$ [$P = 0.0001$] | $lag = 4$ [$P = 0.0456$] |
| Jasper | 28805 | $lag = 2$ [$P = 0.0828$] | $lag = 4$ [$P = 0.0006$] | $lag = 2$ [$P = 0.1565$] |
| Kershaw | 65413 | $lag = 3$ [$P = 0.0008$] | $lag = 1$ [$P = 0.0001$] | $lag = 4$ [$P = 0.0001$] |
| Lancaster | 96012 | $lag = 4$ [$P = 0.0023$] | $lag = 3$ [$P = 0.0001$] | $lag = 3$ [$P = 0.0001$] |
| Lexington | 293912 | $lag = 1$ [$P = 0.0417$] | $lag = 1$ [$P = 0.0001$] | $lag = 3$ [$P = 0.0011$] |
| Marion | 29179 | $lag = 2$ [$P = 0.0011$] | $lag = 4$ [$P = 0.0001$] | $lag = 3$ [$P = 0.0001$] |
| Marlboro | 26669 | $lag = 4$ [$P = 0.1284$] | $lag = 3$ [$P = 0.0210$] | $lag = 1$ [$P = 0.0001$] |
| Richland | 416146 | $lag = 2$ [$P = 0.0036$] | $lag = 3$ [$P = 0.0001$] | $lag = 1$ [$P = 0.0004$] |
| York | 281912 | $lag = 1$ [$P = 0.0460$] | $lag = 4$ [$P = 0.0001$] | $lag = 4$ [$P = 0.0454$] |

**Table S2.** Model parameters and prior distributions used in the spatial Bayesian renewal framework.

| Parameter | Description | Distribution | Explanation |
| --- | --- | --- | --- |
| $g(\tau)$ | the generation interval distribution | $LogNormal(\mu = 3.2, \sigma = 1.64), \quad T = 15$ | Input |
| $d(\tau)$ | the hospital admission delay distribution | $NegBin(\mu = 7, \sigma = 2.49), \quad T = 55$ | Input |
| $\varepsilon(t)$ | Stochastic noise in $R(t)$ | Normal ($\mu=0, \sigma=0.005$) | Estimated |
| $R_0$ | Initial reproductive number | $Truncated\ Normal(R_0^{initial}, scale = 0.1, low = 0)$ | Estimated |
| $\beta_{WW}$ | Wastewater effect coefficient | $Normal(0.01, 0.05)$ | Estimated |
| $\eta$ | Spatial coupling strength | $Normal(0.001, 0.005)$ | Estimated |
| $P_{hosp}$ | Hospitalization probability | $Log\ Normal(\log(2), \log(1.1))$ | Estimated |
| $k_{hosp}$ | Concentration | $Log\ Normal(\log(0.5), \log(10))$ | Estimated |
| $h_i$ | Observed hospital admissions | $Negative\ Binomials(H_i(t), k_{hosp})$ | Likelihood |
| $P_{ij}$ | Normalized mobility probability from county $i$ to county $j$ | Derived from ACS commuting data | Input |

**Table S3.** Comparison of forecasting accuracy (RMSE) for county-level hospital admissions using spatial and non-spatial forecasting models.

|  |  | Forecast COVID |  |  |  |  | Influenza |  |  |  |  | RSV |  |  |  |  |
| --- | --- | --- | --- | --- | --- | --- | --- | --- | --- | --- | --- | --- | --- | --- | --- | --- |
| County Groups | Forecast horizon | AR-IMA | Bayes-Renewal | WW-Bayes-Renewal | Spatial-Bayes-Renewal | Spatial-WW-Bayes-Renewal | AR-IMA | Bayes-Renewal | WW-Bayes-Renewal | Spatial-Bayes-Renewal | Spatial-WW-Bayes-Renewal | AR-IMA | Bayes-Renewal | WW-Bayes-Renewal | Spatial-Bayes-Renewal | Spatial-WW-Bayes-Renewal |
| Sufficient HOSP data + WW data | 1 week | **0.105** | 0.289 | 0.275 | 0.191 | 0.179 | **0.137** | 0.518 | 0.498 | 0.205 | 0.200 | **0.111** | 0.523 | 0.511 | 0.187 | 0.182 |
|  | 2 week | 0.278 | 0.724 | 0.705 | 0.259 | **0.236** | 0.359 | 1.017 | 1.005 | 0.342 | **0.336** | **0.277** | 1.005 | 1.004 | 0.290 | 0.283 |
|  | 3 week | 0.411 | 1.079 | 1.064 | 0.366 | **0.321** | 0.563 | 1.365 | 1.368 | 0.518 | **0.510** | **0.398** | 1.340 | 1.360 | 0.414 | 0.405 |
|  | 4 week | 0.521 | 1.358 | 1.344 | 0.494 | **0.423** | 0.715 | 1.637 | 1.657 | 0.687 | **0.680** | **0.511** | 1.599 | 1.643 | 0.546 | 0.543 |
| Sufficient HOSP data + No WW data | 1 week | **0.105** | 0.294 | - | 0.181 | 0.175 | **0.148** | 0.566 | - | 0.221 | 0.215 | **0.123** | 0.527 | - | 0.211 | 0.210 |
|  | 2 week | 0.278 | 0.743 | - | 0.257 | **0.238** | 0.403 | 1.077 | - | 0.366 | **0.356** | **0.332** | 1.048 | - | 0.341 | 0.332 |
|  | 3 week | 0.403 | 1.105 | - | 0.373 | **0.329** | 0.587 | 1.442 | - | 0.550 | **0.537** | 0.541 | 1.412 | - | 0.470 | **0.454** |
|  | 4 week | 0.531 | 1.381 | - | 0.513 | **0.437** | 0.729 | 1.732 | - | 0.734 | **0.721** | 0.624 | 1.695 | - | 0.604 | **0.580** |
| Limited HOSP data + WW data | 1 week | **0.097** | - | - | 0.466 | 0.113 | **0.136** | - | - | 0.743 | 0.165 | 0.123 | - | - | 0.446 | **0.115** |
|  | 2 week | 0.137 | - | - | 0.497 | **0.131** | **0.201** | - | - | 0.833 | 0.229 | 0.188 | - | - | 0.541 | **0.147** |
|  | 3 week | 0.164 | - | - | 0.552 | **0.149** | **0.240** | - | - | 0.952 | 0.321 | 0.233 | - | - | 0.636 | **0.187** |
|  | 4 week | 0.189 | - | - | 0.633 | **0.197** | **0.247** | - | - | 2.095 | 0.435 | 0.277 | - | - | 0.748 | **0.237** |
| Limited HOSP data + No WW data | 1 week | **0.097** | - | - | 0.410 | 0.117 | **0.134** | - | - | 0.568 | 0.131 | 0.127 | - | - | 0.467 | **0.121** |
|  | 2 week | 0.162 | - | - | 0.427 | **0.139** | 0.206 | - | - | 0.647 | **0.190** | 0.185 | - | - | 0.531 | **0.149** |
|  | 3 week | 0.199 | - | - | 0.463 | **0.169** | **0.270** | - | - | 0.767 | 0.290 | 0.232 | - | - | 0.618 | **0.182** |
|  | 4 week | 0.218 | - | - | 0.523 | **0.210** | **0.314** | - | - | 0.915 | 0.415 | 0.265 | - | - | 0.728 | **0.226** |

**Table S4.** Comparison of forecasting accuracy (WIS) for county-level hospital admissions using spatial and non-spatial forecasting models.

|  |  | Forecast COVID |  |  |  | Influenza |  |  |  | RSV |  |  |  |
| --- | --- | --- | --- | --- | --- | --- | --- | --- | --- | --- | --- | --- | --- |
| County Groups | Forecast horizon | Bayes-Renewal | WW-Bayes-Renewal | Spatial-Bayes-Renewal | Spatial-WW-Bayes-Renewal | Bayes-Renewal | WW-Bayes-Renewal | Spatial-Bayes-Renewal | Spatial-WW-Bayes-Renewal | Bayes-Renewal | WW-Bayes-Renewal | Spatial-Bayes-Renewal | Spatial-WW-Bayes-Renewal |
| Sufficient HOSP data + WW data | 1 week | 3.368 | 3.430 | 6.343 | **1.793** | 6.138 | 7.133 | 7.217 | **2.587** | 2.050 | 2.116 | 2.388 | **1.078** |
|  | 2 week | 4.286 | 4.408 | 6.186 | **2.208** | 8.592 | 10.048 | 7.358 | **6.212** | 2.462 | 2.540 | 2.746 | **1.556** |
|  | 3 week | 5.291 | 5.476 | 6.203 | **3.167** | 11.648 | 13.587 | 9.284 | **13.246** | 2.978 | 3.088 | 3.363 | **2.397** |
|  | 4 week | 6.336 | 6.577 | 6.527 | **4.473** | 15.180 | 17.838 | 13.735 | **23.278** | 3.579 | 3.703 | 4.347 | **3.634** |
| Sufficient HOSP data + No WW data | 1 week | 3.876 | - | 8.984 | **2.085** | 11.290 | - | 17.759 | **3.299** | 2.758 | - | 2.635 | **1.068** |
|  | 2 week | 4.946 | - | 9.145 | **2.733** | 16.560 | - | 18.897 | **7.321** | 3.505 | - | 2.873 | **1.383** |
|  | 3 week | 6.079 | - | 9.748 | **3.918** | 23.208 | - | 22.037 | **15.615** | 4.425 | - | 3.263 | **1.830** |
|  | 4 week | 7.237 | - | 10.832 | **5.485** | 31.255 | - | 28.606 | **27.418** | 5.461 | - | 3.887 | **2.468** |
| Limited HOSP data + WW data | 1 week | - | - | 2.510 | **0.330** | - | - | 6.300 | **0.404** | - | - | 2.462 | **0.198** |
|  | 2 week | - | - | 2.815 | **0.340** | - | - | 7.568 | **0.610** | - | - | 2.801 | **0.216** |
|  | 3 week | - | - | 3.381 | **0.364** | - | - | 10.021 | **1.017** | - | - | 3.334 | **0.236** |
|  | 4 week | - | - | 4.262 | **0.403** | - | - | 14.028 | **1.691** | - | - | 4.161 | **0.271** |
| Limited HOSP data + No WW data | 1 week | - | - | 7.799 | **0.440** | - | - | 10.711 | **0.404** | - | - | 2.939 | **0.197** |
|  | 2 week | - | - | 8.309 | **0.457** | - | - | 12.526 | **0.589** | - | - | 3.340 | **0.217** |
|  | 3 week | - | - | 9.322 | **0.494** | - | - | 16.144 | **1.002** | - | - | 3.963 | **0.240** |
|  | 4 week | - | - | 11.045 | **0.554** | - | - | 22.147 | **1.708** | - | - | 4.941 | **0.276** |

**Figure S1.**
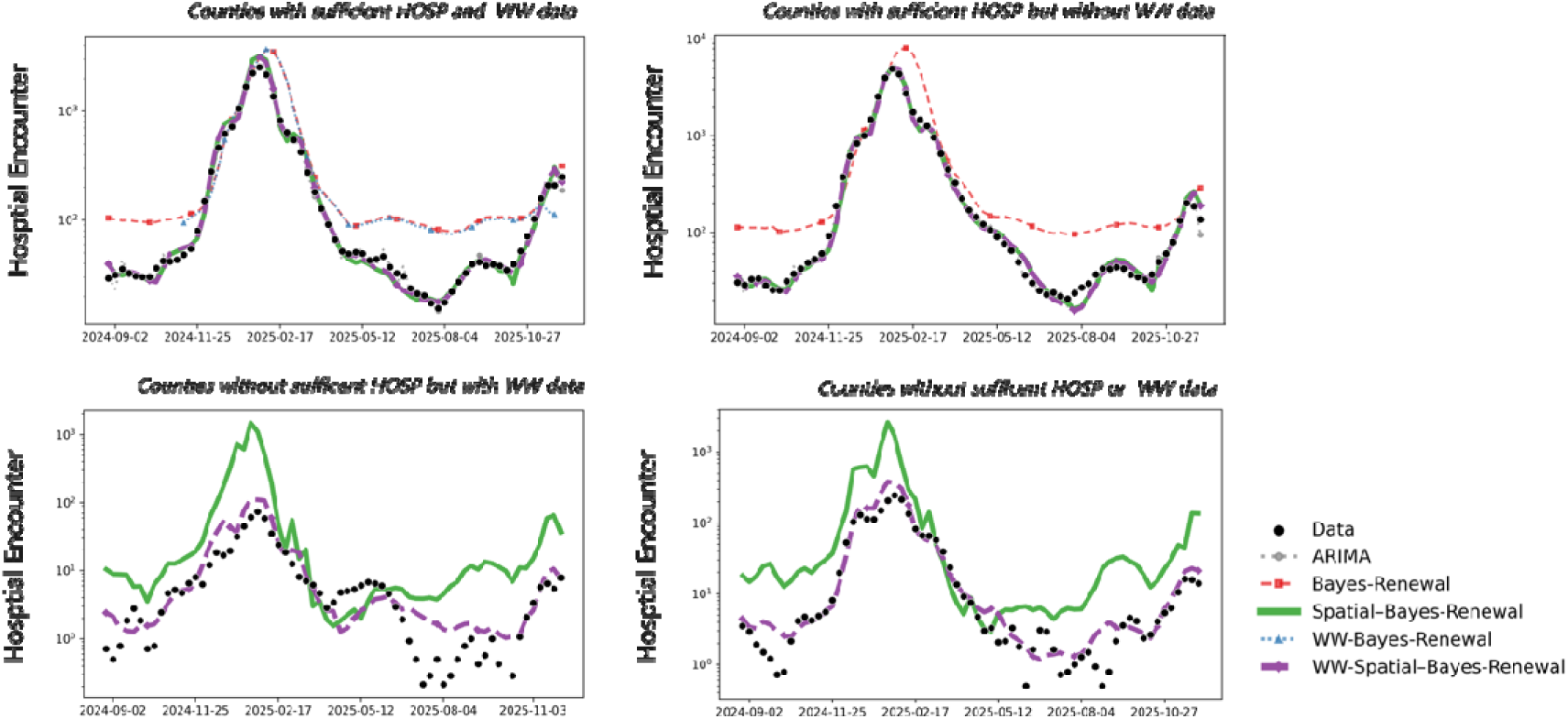
County-level forecasts of Influenza hospital admissions under different surveillance settings.

**Figure S2.**
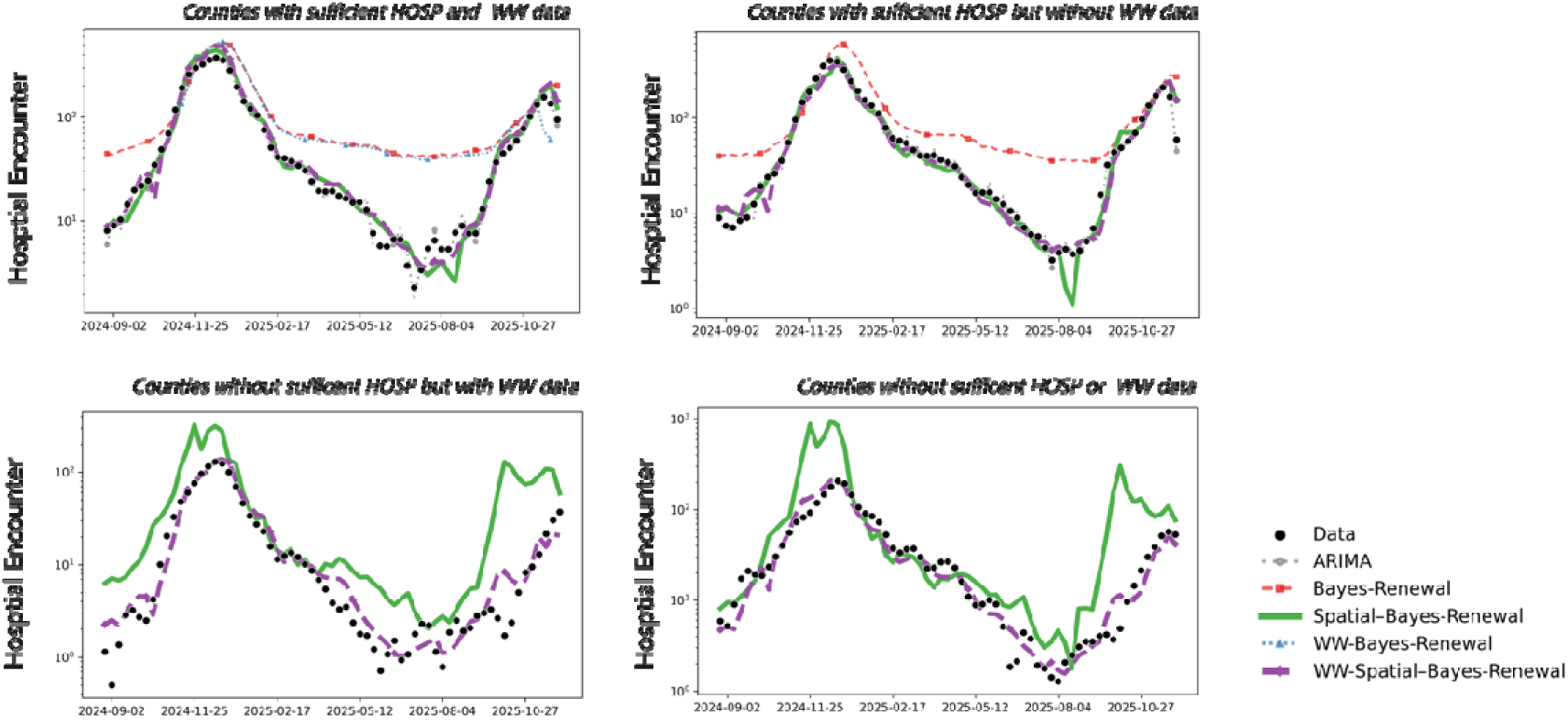
County-level forecasts of RSV hospital admissions under different surveillance settings.

**Figure S3.**
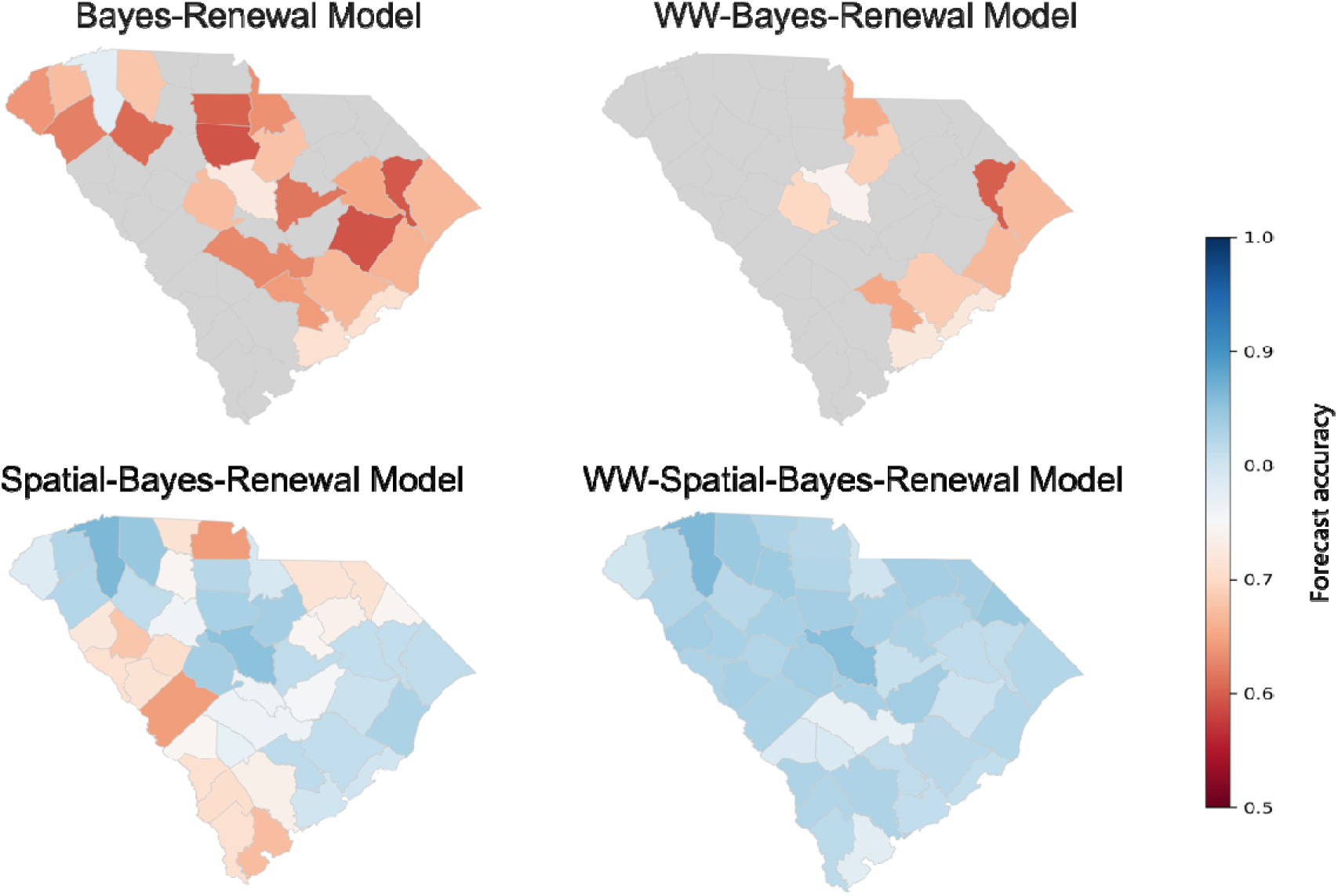
Spatial variation of forecast accuracy for Influenza across SC counties.

**Figure S4.**
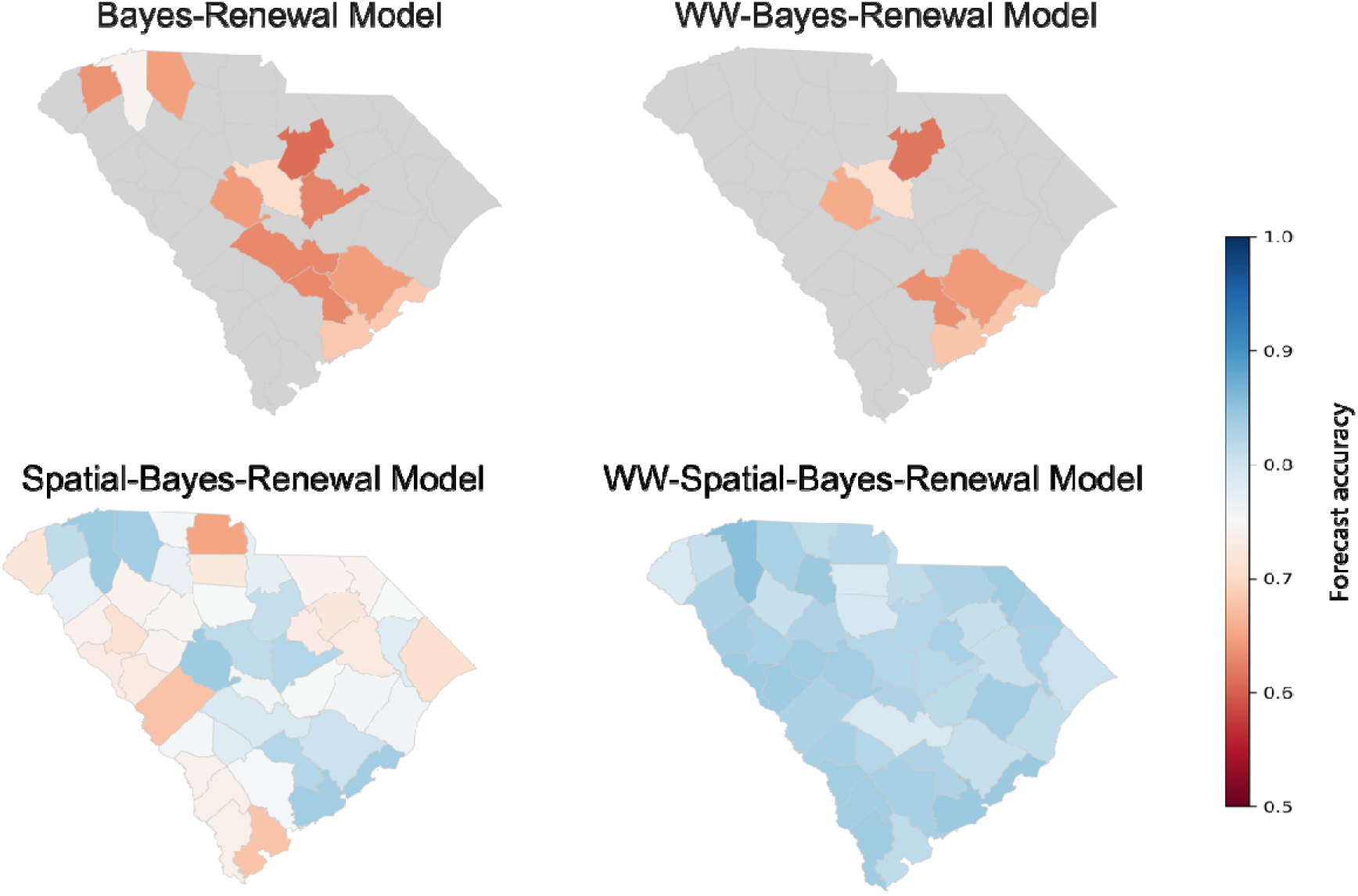
Spatial variation of forecast accuracy for RSV across SC counties.

## Notes

### Competing Interest Statement

The authors have declared no competing interest.

### Author Declarations

The data "county-level and facility-level hospital admission data for respiratory illnesses" used in our study were aggregated/summary data.

### Summary of Updates

Section on Abstract, Introduction, Discussion has been revised.

